# How is advocacy defined, conceptualised and implemented within nursing, midwifery and the allied health professions? A protocol for a systematic review of the evidence

**DOI:** 10.1101/2025.04.08.25325333

**Authors:** Sara Tavares, Isla Kuhn, Mary E. Harrison, Gemma Caughers, Juan Carlos Quijano-Campos, Joana Oliveira, Irene Gibson, Donna Fitzsimons, Ali Ghanchi, Peter Hartley, Marie Friedel, Faye Forsyth

## Abstract

**Aim:** To explore how advocacy has been defined, conceptualised and implemented within nursing, midwifery and the allied health professions. A secondary aim is to characterize barriers and facilitators to advocacy in clinical practice.

**Methods:** A systematic search of five bibliographic databases has been undertaken to identify primary qualitative and quantitative research relating to advocacy in nursing, midwifery and the allied health professions. Results from searches are being screened blinded and in duplicate. Given that we anticipate retrieving heterogenous studies, eligible studies will be synthesised according to best practice set out in a range of guidelines.

**Data Sources:** CINAHL (via EBSCOHost), EMBASE (via Ovid), MEDLINE (via Ovid), PsycINFO (via EBSCOHost), Global Health (via EBSCOHost) have been searched.

**Results:** Searches have been completed, n=45,617 studies were retrieved which was reduced to n=25,900 studies following deduplication. Screening, which is being performed blinded and in duplicate within the web platform Rayyan, is ongoing.

**Conclusions:** As a result of this robust review, we anticipate generating a comprehensive report of advocacy that we have hitherto identified as being lacking. Best practice principles are being employed ensuring the eventual output is robust, transparent and useful to the clinicians likely to be accessing this information.

**No Patient or Public Contribution:** *Implications for the profession and/or patient care:* In order to gain a comprehensive understanding of advocacy; nurses, midwives and allied health professionals would have to obtain and consult a number of systematic reviews. This all-encompassing review will ensure that a comprehensive report that synthesizes modern data and covers the full-spectrum of advocacy domains and its implications is available.

*Impact:* The broad reach and ambitious nature of this review will ensure it has a significant impact. Advocacy is practiced by most healthcare staff on daily basis, however many are not aware of this and those with aspirations to engage more fully as an advocate within their role do not have a reliable and comprehensive report which would equip them with relevant knowledge relating to advocacy.

*Protocol Registration:* This review was registered on PROSPERO (CRD420250640543)

*Key points:*

- Advocacy is an important component of nursing, midwifery and allied health professional practice, however the evidence has not recently been synthesised.
- This review will update and expand the scope of previous reviews, to generate a comprehensive update of advocacy.
- Clinicians with an interest in advocacy will be able to obtain a robust and up to date overview of advocacy, which is likely to have changed recently in view of the spread of misinformation and artificial intelligence.

*Registration:* This review was registered on PROSPERO (CRD420250640543)

## Introduction

Advocacy is a complex concept that remains ill-defined and under theorised.^1^ Despite this, it is widely regarded as a key component of professional practice,^2^ and it features in the majority of codes of practice within nursing, midwifery and the allied health professions.^3,4^ Broadly speaking, advocacy is perceived to be a critical mechanism through which the healthcare workforce can advance and uphold their professions, improve patient care and outcomes, and influence healthcare policy.^5^ However, the conceptualisation and practice of advocacy is highly variable,^6^ therefore most reviews have limited their remit to synthesizing one component such as definitions,^7,8^ conceptualisations,^6,9^ or practice.^10,11^ To our knowledge, no previous review has adopted a wide lens and attempted to comprehensively summarise the multiple threads of advocacy.

## Rationale

Whilst there are several reviews examining advocacy in nursing and the allied health professions,^6-11^ none have simultaneously synthesised the key tenets of definition, conceptualisation, implementation and barriers and facilitators, as proposed here. Consequently, a broad overview of advocacy in nursing and the allied health professions cannot be obtained without first visiting multiple publications. In addition, there has been unprecedented global events (COVID-19, infodemics and misinformation)^12^ and developments in information technology (artificial intelligence),^13^ which have the potential to change how we perceive advocacy and how it is implemented in clinical practice.^14,15^ Therefore, an all-encompassing review that updates previous efforts and which culminates in the production of a single comprehensive report is warranted.

## Objectives

This review has four objectives that aim to establish:

1. how advocacy has been defined across the nursing, midwifery and allied health professional literature;
2. how advocacy has been conceptualised (domains, categories, levels) within the nursing and allied health professional literature;
3. how advocacy has been implemented by nurses and allied health professionals working in the various domains of advocacy;
4. the barriers and facilitators to advocacy described by nurses and allied health professionals.

## Methods

This report has been structured in line with PRISMA-P (Preferred Reporting Items for Systematic review and Meta-Analysis Protocols) guidance.^16^

### Eligibility criteria

The PICO (population, intervention, comparison, outcome) framework^17^ has been used to structure the eligibility criteria. We will include any type of primary research (i.e. qualitative, quantitative) as long as they meet inclusion criteria. We will exclude editorials, opinion pieces or conference abstracts as they do not undergo the same level of rigorous peer review. We will not include systematic reviews, all prior systematic reviews on advocacy will be located and a hand search of the reference list undertaken. Only studies published in the English language will be considered owing to the composition of the team undertaken screening and extraction. No date limits have been set as the intention is to capture any evolution in definitions, conceptualisation and implementation. Allied health professional search criteria has been derived from the United Kingdom, as they are one of the few large bodies to provide a definition. However, equivalent job titles will be considered during the screening process.

**Table 1.** PICO criteria.

| Criterion | Description |
| --- | --- |
| Population | Nurses, midwives and allied health professionals.* Studies with mixed |
|  | professionals groups will be considered and included if there is sufficient description of the sample to determine the proportion of nurses, midwives, or allied health professionals. Healthcare assistants will not be included as they do fall under the key terms of nurses, midwives and allied health professionals. |
| **Intervention** | Advocacy and related terms (for example: peer advocacy, self-advocacy, empowerment, patient representative, patient activism) that are discussed within any adult healthcare and healthcare related setting (i.e. community, healthcare policy, funding and policy making, social research and education). Definitions of adult vary country by country, therefore each study will be assessed and included only if the advocacy activities described are performed in adult care settings. |
| **Comparison** | Not applicable |
| **Outcome** | Definitions of advocacy, conceptualisations of advocacy, descriptions of the operationalisation of advocacy in healthcare practice, barriers and facilitators to implementing advocacy in practice. |

\*the National Health Service in England recognises 14 different allied health professions and the National Institute of Health and Care Research defines allied health professionals as all health and care professionals excluding doctors and dentists. These definitions will guide search terms.<sup>18</sup>

### Information sources

Five databases CINAHL (via EBSCOHost), EMBASE (via Ovid), MEDLINE (via Ovid), PsycINFO (via EBSCOHost), Global Health (via EBSCOHost) have been searched. These databases were selected on the basis of pilot search results. As per recommendations, we will review the reference lists of included studies and relevant contemporary systematic reviews to ensure comprehensive data capture.^16^ The search will be updated before submission to a peer reviewed journal for publication.

### Search strategy

Search terms have been developed and piloted in conjunction with an experienced information specialist (IK). Databases were searched separately and search syntaxes were adapted to the respective databases, in line with best practice.^19^ Search results were deduplicated in Endnote by IK, using established methods.^20^ Before finalising, the search strategy was assessed against the PRISMA-S (Preferred Reporting Items for Systematic reviews and Meta-Analyses literature search extension) guidelines.^19^ Searches included the period from database inception until present. The full searches can be viewed in the Supplementary Material.

### Study records

#### Data management

Data will be managed using the following platforms Endnote^21^ (search results and deduplication), Rayyan^22^ (screening), Microsoft Excel^23^ and Nvivo^24^ (data extraction).

#### Selection process

Two blinded reviewers will independently evaluate all titles and abstracts against the inclusion criteria within the platform Rayyan. Reviewers will assign one of the following labels to each search result: include, exclude, maybe. Studies labelled include and maybe will undergo full-text review, which will also be performed blinded and in duplicate. Where agreement cannot be reached, we will seek additional information from the study authors and/or consult a third party to adjudicate. Reasons for exclusion will be recorded and documented within the final report.

#### Data collection process

Data extraction proformas will be piloted prior to use. One author will extract data, ten percent of extracted data will be checked by a second author. Any disagreements will be resolved through discussion. We will extract data within the following domains: administrative, demographic, textual descriptions of results, empirical outcomes, conclusions. Data extraction proformas may be modified during the extraction process, version control methods will be employed to document changes to proformas.^25^

#### Data items

Within the administrative domain we will extract the following data points: title, authors, date, journal, country, design, methods. The demographic fields that will be extracted include: number of participants, types of participants, categorisation of professionals, seniority of professionals. Textual descriptions of data will be abstracted using NVivo software^24^ to enable documentation and characterisation of definitions, conceptualisations, and activities. Outcome data, for example results from surveys, will be tabulated within Microsoft Excel software and described using the appropriate statistical methods.

#### Outcomes and prioritization

Our primary outcomes are definitions of advocacy, conceptualisations of advocacy, descriptions of advocacy activities, descriptions of facilitators, descriptions of barriers. Descriptions will be abstracted in full, however, we will apply categorisations and counts if the data is deemed amenable. For example, if advocacy is conceptualised differently by two different papers, we will attempt to distill the components they include within the conceptualisation to provide a count and overview of the data in full.

#### Risk of bias in individual studies

We anticipate retrieving qualitative, quantitative, and mixed methods studies, therefore, we have elected to use the Mixed Methods Appraisal Tool (MMAT) version 2018 for information professionals and researchers^26^ for risk of bias assessment. The assessment will be performed by one author, ten percent will be assessed blind and in duplicate. Divergences in arbitrations will be resolved through discussion, a third party will be consulted if necessary.

### Data synthesis

Pilot searches have retrieved studies with predominantly qualitative designs. However, we have identified quantitative and mixed methods studies that are potentially eligible via preliminary reference checks (surveys or combined interview and survey studies). Therefore, we will extract different types of data separately (e.g. qualitative and quantitative data), and then integrate these sequentially in a narrative synthesis following Joanna Briggs Institute guidance outlined by Stern et al.^27^ Definitions and conceptualisations of advocacy will most likely be text data, these will be extracted in their text data format and interrogated for their components to enable comparison and counting. Descriptions of how advocacy is implemented and barriers and facilitators will also be textual, these data will be analysed thematically within the platform Nvivo to determine commonalities/divergence. We do not anticipate using statistical analysis methods such as meta-analysis given the type of data likely to be retrieved, however we will use counts and ranges as required.

## Conclusion

This review aims to provide a comprehensive report on advocacy in nursing and the allied professions. Currently, professionals in either discipline would have to visit multiple reports to generate such an expansive overview, further, some of the reports available may not capture contemporary insights. A review that employs robust methods and transparent reporting, as outlined here, is therefore necessary to address this research need.

## Relevance for clinical practice

Advocacy has likely changed in clinical practice as a result of advances in artificial intelligence and the increasing prevalence of misinformation. Clinicians have a duty of care to be aware of such changes and respond accordingly. This review of advocacy, which updates and expands on previous reviews, will provide clinicians with a robust and comprehensive report which they can use to enhance their knowledge and improve they practice advocacy in the clinical setting.

## Data Availability

All relevant data from this study will be made available upon study completion.

