## Supplementary material for "How is advocacy defined, conceptualised and implemented within nursing, midwifery and the allied health professions? A protocol for a systematic review of the evidence": Supplemantary data

### Overview of searches performed on 25 Feb 2025

| Medline via ovid | 12,100 |
| --- | --- |
| Embase via OVID | 14,563 |
| CINAHL via EbscoHost | 13,781 |
| PsycINFO via EbscoHost | 4,211 |
| Global Health via EbscoHost | 962 |
| **Total** | **45,617** |
| **Total deduplicated** | **25,900** |

### Medline

Ovid MEDLINE(R) and Epub Ahead of Print, In-Process, In-Data-Review & Other Non-Indexed Citations, Daily and Versions <1946 to February 24, 2025>

<https://ovidsp.ovid.com/athens/ovidweb.cgi?T=JS&NEWS=N&PAGE=main&SHAREDSEARCHID=JbamNhvWE5R0K5b1pMxouJpPznLyLBlWIP7ykGBpRXrHrvCCdg7WhLcJy8LoGoOR>

1	(Nurse* or allied health professional* or allied professional* or art therapist* or dietitian* or drama therapist* or music therapist* or occupational therapist* or operating department practitioner* or orthoptist* or osteopath* or paramedic* or physiotherapist* or podiatrist* or prosthetist* or orthotist* or radiographer* or "speech and language therapist*" or "speech therapist*").ti,ab,kw,kf.	399831

2	exp Nurses/ or allied health personnel/ or exp paramedics/ or exp physical therapist assistants/ or exp occupational therapists/ or exp physical therapists/ or exp Nutritionists/ or exp Osteopathic Physicians/	122064

3	1 or 2	453413

4	(advocacy or advocat* or selfadvocacy or "self advocat*" or empower* or "speak* out" or "whistelblow*" or "whistle blow*" or activist* or activism or campaigner* or ombudsman*).ti,ab,kw,kf.	158589

5	exp Patient Advocacy/ or exp Empowerment/ or exp Political Activism/	25976

6	exp Whistleblowing/	1248

7	4 or 5 or 6	179550

8	(defin* or concept* or evaluat* or domain* or categor* or operationali* or implement* or taxonom* or barrier* or role* or activity or activities or experience* or facilitator).ti,ab,kw,kf.	14361595

9	(qualitative or survey or cross-sectional or longitudinal).ti,ab,kw,kf.	1894451

10	exp Qualitative Research/	99968

11	exp "Surveys and Questionnaires"/	1295033

12	exp Cross-Sectional Studies/	533319

13	exp Longitudinal Studies/	181116

14	exp Nurse's Role/ or exp Professional Role/ or exp Role/	115522

15	exp Classification/	274092

16	8 or 9 or 10 or 11 or 12 or 13 or 14 or 15	15805744

17	3 and 7 and 16	12100

### Embase

Embase <1974 to 2025 February 24>

https://ovidsp.ovid.com/athens/ovidweb.cgi?T=JS&NEWS=N&PAGE=main&SHAREDSEARCHID=7cwAQf6plFGfKUItnnEHmqXiobnmemmqzoZDhgrT0KU37E9XpEHFC3hc47gFjK8LW

1	(Nurse* or allied health professional* or allied professional* or art therapist* or dietitian* or drama therapist* or music therapist* or occupational therapist* or operating department practitioner* or orthoptist* or osteopath* or paramedic* or physiotherapist* or podiatrist* or prosthetist* or orthotist* or radiographer* or "speech and language therapist*" or "speech therapist*").ti,ab.	492227

2	exp *nurse/	90454

3	exp *eye care professional/ or exp *orthotist/ or exp *prosthetist/	2281

4	exp *dietitian/	2391

5	paramedical personnel/ or exp *audiologist/ or exp *nursing staff/ or exp *occupational therapist/ or exp *operating room personnel/ or exp *physiotherapist/ or exp *radiographer/ or exp *speech language pathologist/	75814

6	exp *orthoptist/	31

7	exp *osteopathic physician/	185

8	exp *rescue personnel/	4700

9	exp *podiatrist/	90

10	or/1-9	568080

11	(advocacy or advocat* or selfadvocacy or "self advocat*" or empower* or "speak* out" or "whistelblow*" or "whistle blow*" or activist* or activism or campaigner* or ombudsman*).ti,ab.	201245

12	exp *patient advocacy/ or exp *advocacy group/ or exp *self advocacy/	11559

13	exp *patient empowerment/ or exp *empowerment/	3813

14	exp *activism/	11511

15	exp *whistleblowing/	774

16	or/11-15	210668

17	(defin* or concept* or evaluat* or domain* or categor* or operationali* or implement* or taxonom* or barrier* or role* or activity or activities or experience* or facilitator).ti,ab.	18242655

18	(qualitative or survey or cross-sectional or longitudinal).ti,ab.	2404660

19	exp *concept analysis/ or exp *concept mapping/	941

20	exp *taxonomy/	139833

21	exp *qualitative research/ or exp *qualitative analysis/	32313

22	exp *questionnaire/	48164

23	exp *cross-sectional study/	14592

24	exp *short survey/	76

25	exp *longitudinal study/	9334

26	or/17-25	19225085

27	10 and 16 and 26	14563

### PsycINFO

| **#** | **Query** | **Limiters/Expanders** | **Last Run Via** | **Results** |
| --- | --- | --- | --- | --- |
| S1 | TI(Nurse* or “allied health professional”* or “allied professional*” or “art therapist*” or dietitian* or “drama therapist*” or “music therapist*” or “occupational therapist*” or “operating department practitioner*” or orthoptist* or osteopath* or paramedic* or physiotherapist* or podiatrist* or prosthetist* or orthotist* or radiographer* or "speech and language therapist*" or "speech therapist*") or AB(Nurse* or “allied health professional”* or “allied professional*” or “art therapist*” or dietitian* or “drama therapist*” or “music therapist*” or “occupational therapist*” or “operating department practitioner*” or orthoptist* or osteopath* or paramedic* or physiotherapist* or podiatrist* or prosthetist* or orthotist* or radiographer* or "speech and language therapist*" or "speech therapist*") | Expanders - Apply equivalent subjects Search modes - Proximity | Interface - EBSCOhost Research Databases Search Screen - Basic Search Database - APA PsycInfo | 98,625 |
| S2 | DE "Nurses" OR DE "Nurse Practitioners" OR DE "Psychiatric Nurses" OR DE "Public Health Service Nurses" OR DE "School Nurses" | Expanders - Apply equivalent subjects Search modes - Proximity | Interface - EBSCOhost Research Databases Search Screen - Basic Search Database - APA PsycInfo | 43,857 |
| S3 | (DE "Allied Health Personnel") OR (DE "Occupational Therapists" OR DE "Physical Therapists" OR DE "Speech Therapists") | Expanders - Apply equivalent subjects Search modes - Proximity | Interface - EBSCOhost Research Databases Search Screen - Basic Search Database - APA PsycInfo | 8,066 |
| S4 | (DE "Nutritionists") | Expanders - Apply equivalent subjects Search modes - Proximity | Interface - EBSCOhost Research Databases Search Screen - Basic Search Database - APA PsycInfo | 114 |
| S5 | (DE "Paramedics") OR (DE "Speech Therapists") | Expanders - Apply equivalent subjects Search modes - Proximity | Interface - EBSCOhost Research Databases Search Screen - Basic Search Database - APA PsycInfo | 2,505 |
| S6 | S1 OR S2 OR S3 OR S4 OR S5 | Expanders - Apply equivalent subjects Search modes - Proximity | Interface - EBSCOhost Research Databases Search Screen - Basic Search Database - APA PsycInfo | 106,722 |
| S7 | TI (advocacy or advocat* or selfadvocacy or "self advocat*" or empower* or "speak* out" or "whistelblow*" or "whistle blow*" or activist* or activism or campaigner* or ombudsman*) or AB (advocacy or advocat* or selfadvocacy or "self advocat*" or empower* or "speak* out" or "whistelblow*" or "whistle blow*" or activist* or activism or campaigner* or ombudsman*) | Expanders - Apply equivalent subjects Search modes - Proximity | Interface - EBSCOhost Research Databases Search Screen - Basic Search Database - APA PsycInfo | 107,411 |
| S8 | (((DE "Advocacy" OR DE "Community Advocacy" OR DE "Self-Advocacy") OR (DE "Empowerment")) OR (DE "Informants")) OR (DE "Activism" OR DE "Student Activism") | Expanders - Apply equivalent subjects Search modes - Proximity | Interface - EBSCOhost Research Databases Search Screen - Basic Search Database - APA PsycInfo | 26,412 |
| S9 | S7 OR S8 | Expanders - Apply equivalent subjects Search modes - Proximity | Interface - EBSCOhost Research Databases Search Screen - Basic Search Database - APA PsycInfo | 111,312 |
| S10 | TI(defin* or concept* or evaluat* or domain* or categor* or operationali* or implement* or taxonom* or barrier* or role* or activity or activities or experience* or facilitator) or AB (defin* or concept* or evaluat* or domain* or categor* or operationali* or implement* or taxonom* or barrier* or role* or activity or activities or experience* or facilitator) | Expanders - Apply equivalent subjects Search modes - Proximity | Interface - EBSCOhost Research Databases Search Screen - Basic Search Database - APA PsycInfo | 3,023,047 |
| S11 | TI(qualitative or survey or cross-sectional or longitudinal) or AB (qualitative or survey or cross-sectional or longitudinal) | Expanders - Apply equivalent subjects Search modes - Proximity | Interface - EBSCOhost Research Databases Search Screen - Basic Search Database - APA PsycInfo | 788,607 |
| S12 | ((DE "Concept Formation" OR DE "Cognitive Discrimination" OR DE "Cognitive Generalization") ) OR (DE "Professional Role" OR DE "Counselor Role" OR DE "Professional Boundaries" OR DE "Therapist Role") | Expanders - Apply equivalent subjects Search modes - Proximity | Interface - EBSCOhost Research Databases Search Screen - Basic Search Database - APA PsycInfo | 33,606 |
| S13 | ((DE "Qualitative Measures" OR DE "Diary Measure" OR DE "Semi-Structured Interview" OR DE "Qualitative Methods" OR DE "Coding Scheme" OR DE "Content Analysis" OR DE "Ethnography" OR DE "Focus Group" OR DE "Grounded Theory" OR DE "Interpretative Phenomenological Analysis" OR DE "Narrative Analysis" OR DE "Semi-Structured Interview" OR DE "Thematic Analysis") OR (DE "Surveys" OR DE "Consumer Surveys" OR DE "Mail Surveys" OR DE "Online Surveys" OR DE "Telephone Surveys")) OR (DE "Longitudinal Studies" OR DE "Prospective Studies") | Expanders - Apply equivalent subjects Search modes - Proximity | Interface - EBSCOhost Research Databases Search Screen - Basic Search Database - APA PsycInfo | 145,694 |
| S14 | S10 OR S11 OR S12 OR S13 | Expanders - Apply equivalent subjects Search modes - Proximity | Interface - EBSCOhost Research Databases Search Screen - Basic Search Database - APA PsycInfo | 3,330,763 |
| S15 | S6 AND S9 AND S14 | Expanders - Apply equivalent subjects Search modes - Proximity | Interface - EBSCOhost Research Databases Search Screen - Basic Search Database - APA PsycInfo | 4,211 |

### Global Health

| **#** | **Query** | **Limiters/Expanders** | **Last Run Via** | **Results** |
| --- | --- | --- | --- | --- |
| S1 | TI(Nurse* or “allied health professional”* or “allied professional*” or “art therapist*” or dietitian* or “drama therapist*” or “music therapist*” or “occupational therapist*” or “operating department practitioner*” or orthoptist* or osteopath* or paramedic* or physiotherapist* or podiatrist* or prosthetist* or orthotist* or radiographer* or "speech and language therapist*" or "speech therapist*") or AB(Nurse* or “allied health professional”* or “allied professional*” or “art therapist*” or dietitian* or “drama therapist*” or “music therapist*” or “occupational therapist*” or “operating department practitioner*” or orthoptist* or osteopath* or paramedic* or physiotherapist* or podiatrist* or prosthetist* or orthotist* or radiographer* or "speech and language therapist*" or "speech therapist*") | Expanders - Apply equivalent subjects Search modes - Proximity | Interface - EBSCOhost Research Databases Search Screen - Basic Search Database - Global Health | 42,513 |
| S2 | TI (advocacy or advocat* or selfadvocacy or "self advocat*" or empower* or "speak* out" or "whistelblow*" or "whistle blow*" or activist* or activism or campaigner* or ombudsman*) or AB (advocacy or advocat* or selfadvocacy or "self advocat*" or empower* or "speak* out" or "whistelblow*" or "whistle blow*" or activist* or activism or campaigner* or ombudsman*) | Expanders - Apply equivalent subjects Search modes - Proximity | Interface - EBSCOhost Research Databases Search Screen - Basic Search Database - Global Health | 31,974 |
| S3 | TI(defin* or concept* or evaluat* or domain* or categor* or operationali* or implement* or taxonom* or barrier* or role* or activity or activities or experience* or facilitator) or AB (defin* or concept* or evaluat* or domain* or categor* or operationali* or implement* or taxonom* or barrier* or role* or activity or activities or experience* or facilitator) | Expanders - Apply equivalent subjects Search modes - Proximity | Interface - EBSCOhost Research Databases Search Screen - Basic Search Database - Global Health | 2,269,495 |
| S4 | TI(qualitative or survey or cross-sectional or longitudinal) or AB (qualitative or survey or cross-sectional or longitudinal) | Expanders - Apply equivalent subjects Search modes - Proximity | Interface - EBSCOhost Research Databases Search Screen - Basic Search Database - Global Health | 558,196 |
| S5 | S3 OR S4 | Expanders - Apply equivalent subjects Search modes - Proximity | Interface - EBSCOhost Research Databases Search Screen - Basic Search Database - Global Health | 2,516,007 |
| S6 | S1 AND S2 AND S5 | Expanders - Apply equivalent subjects Search modes - Proximity | Interface - EBSCOhost Research Databases Search Screen - Basic Search Database - Global Health | 962 |

### CINAHL

| **#** | **Query** | **Limiters/Expanders** | **Last Run Via** | **Results** |
| --- | --- | --- | --- | --- |
| S1 | TI(Nurse* or “allied health professional”* or “allied professional*” or “art therapist*” or dietitian* or “drama therapist*” or “music therapist*” or “occupational therapist*” or “operating department practitioner*” or orthoptist* or osteopath* or paramedic* or physiotherapist* or podiatrist* or prosthetist* or orthotist* or radiographer* or "speech and language therapist*" or "speech therapist*") or AB(Nurse* or “allied health professional”* or “allied professional*” or “art therapist*” or dietitian* or “drama therapist*” or “music therapist*” or “occupational therapist*” or “operating department practitioner*” or orthoptist* or osteopath* or paramedic* or physiotherapist* or podiatrist* or prosthetist* or orthotist* or radiographer* or "speech and language therapist*" or "speech therapist*") | Expanders - Apply equivalent subjects  Search modes - Proximity | Interface - EBSCOhost Research Databases  Search Screen - Advanced Search  Database - CINAHL | 415,407 |
| S2 | (MH "Nurses+") OR (MH "Operating Room Personnel+") OR (MH "Podiatrists") OR (MH "Dietitians") OR (MH "Paramedics") OR (MH "Ophthalmic Technologists") OR (MH "Operating Department Practitioners") OR (MH "Nutritionists") OR (MH "Physical Therapists+") OR (MH "Speech-Language Pathologists") OR (MH "Occupational Therapists+") OR (MH "Music Therapists") OR (MH "Art Therapists") OR (MH "Osteopaths") OR (MH "Radiologic Technologists+") | Expanders - Apply equivalent subjects  Search modes - Proximity | Interface - EBSCOhost Research Databases  Search Screen - Advanced Search  Database - CINAHL | 311,172 |
| S3 | S1 OR S2 | Expanders - Apply equivalent subjects  Search modes - Proximity | Interface - EBSCOhost Research Databases  Search Screen - Advanced Search  Database - CINAHL | 585,008 |
| S4 | TI (advocacy or advocat* or selfadvocacy or "self advocat*" or empower* or "speak* out" or "whistelblow*" or "whistle blow*" or activist* or activism or campaigner* or ombudsman*) or AB (advocacy or advocat* or selfadvocacy or "self advocat*" or empower* or "speak* out" or "whistelblow*" or "whistle blow*" or activist* or activism or campaigner* or ombudsman*) | Expanders - Apply equivalent subjects  Search modes - Proximity | Interface - EBSCOhost Research Databases  Search Screen - Advanced Search  Database - CINAHL | 84,658 |
| S5 | (MH "Patient Advocacy") OR (MH "Self-Advocacy") OR (MH "Whistle Blowing") OR (MH "Political Participation") | Expanders - Apply equivalent subjects  Search modes - Proximity | Interface - EBSCOhost Research Databases  Search Screen - Advanced Search  Database - CINAHL | 32,460 |
| S6 | S4 OR S5 | Expanders - Apply equivalent subjects  Search modes - Proximity | Interface - EBSCOhost Research Databases  Search Screen - Advanced Search  Database - CINAHL | 108,788 |
| S7 | TI(defin* or concept* or evaluat* or domain* or categor* or operationali* or implement* or taxonom* or barrier* or role* or activity or activities or experience* or facilitator) or AB (defin* or concept* or evaluat* or domain* or categor* or operationali* or implement* or taxonom* or barrier* or role* or activity or activities or experience* or facilitator) | Expanders - Apply equivalent subjects  Search modes - Proximity | Interface - EBSCOhost Research Databases  Search Screen - Advanced Search  Database - CINAHL | 2,602,968 |
| S8 | TI(qualitative or survey or cross-sectional or longitudinal) or AB (qualitative or survey or cross-sectional or longitudinal) | Expanders - Apply equivalent subjects  Search modes - Proximity | Interface - EBSCOhost Research Databases  Search Screen - Advanced Search  Database - CINAHL | 779,459 |
| S9 | (MH "Qualitative Studies+") OR (MH "Cross Sectional Studies") OR (MH "Surveys+") OR (MH "Prospective Studies+") | Expanders - Apply equivalent subjects  Search modes - Proximity | Interface - EBSCOhost Research Databases  Search Screen - Advanced Search  Database - CINAHL | 1,184,920 |
| S10 | (MH "Concept Formation") OR (MH "Concept Mapping") OR (MH "Concept Analysis") OR (MH "Classification+") OR (MH "Role+") OR (MH "Professional Role+") | Expanders - Apply equivalent subjects  Search modes - Proximity | Interface - EBSCOhost Research Databases  Search Screen - Advanced Search  Database - CINAHL | 210,147 |
| S11 | S7 OR S8 OR S9 OR S10 | Expanders - Apply equivalent subjects  Search modes - Proximity | Interface - EBSCOhost Research Databases  Search Screen - Advanced Search  Database - CINAHL | 3,384,689 |
| S12 | S3 AND S6 AND S11 | Expanders - Apply equivalent subjects  Search modes - Proximity | Interface - EBSCOhost Research Databases  Search Screen - Advanced Search  Database - CINAHL | 13,781 |
